# Can we trust the prediction model? Demonstrating the importance of external validation by investigating the COVID-19 Vulnerability (C-19) Index across an international network of observational healthcare datasets

**DOI:** 10.1101/2020.06.15.20130328

**Authors:** Jenna M. Reps, Chungsoo Kim, Ross D. Williams, Aniek F. Markus, Cynthia Yang, Talita Duarte Salles, Thomas Falconer, Jitendra Jonnagaddala, Andrew Williams, Sergio Fernández-Bertolín, Scott L. DuVall, Kristin Kostka, Gowtham Rao, Azza Shoaibi, Anna Ostropolets, Matthew E Spotnitz, Lin Zhang, Paula Casajust, Ewout W. Steyerberg, Fredrik Nyberg, Benjamin Skov Kaas-Hansen, Young Hwa Choi, Daniel Morales, Siaw-Teng Liaw, Maria Tereza Fernandes Abrahão, Carlos Areia, Michael E. Matheny, María Aragón, Rae Woong Park, George Hripcsak, Christian G. Reich, Marc A. Suchard, Seng Chan You, Patrick B. Ryan, Daniel Prieto-Alhambra, Peter R. Rijnbeek

## Abstract

**Background:** SARS-CoV-2 is straining healthcare systems globally. The burden on hospitals during the pandemic could be reduced by implementing prediction models that can discriminate between patients requiring hospitalization and those who do not. The COVID-19 vulnerability (C-19) index, a model that predicts which patients will be admitted to hospital for treatment of pneumonia or pneumonia proxies, has been developed and proposed as a valuable tool for decision making during the pandemic. However, the model is at high risk of bias according to the Prediction model Risk Of Bias ASsessment Tool and has not been externally validated.

**Methods:** We followed the OHDSI framework for external validation to assess the reliability of the C-19 model. We evaluated the model on two different target populations: i) 41,381 patients that have SARS-CoV-2 at an outpatient or emergency room visit and ii) 9,429,285 patients that have influenza or related symptoms during an outpatient or emergency room visit, to predict their risk of hospitalization with pneumonia during the following 0 to 30 days. In total we validated the model across a network of 14 databases spanning the US, Europe, Australia and Asia.

**Findings:** The internal validation performance of the C-19 index was a c-statistic of 0.73 and calibration was not reported by the authors. When we externally validated it by transporting it to SARS-CoV-2 data the model obtained c-statistics of 0.36, 0.53 (0.473-0.584) and 0.56 (0.488-0.636) on Spanish, US and South Korean datasets respectively. The calibration was poor with the model under-estimating risk. When validated on 12 datasets containing influenza patients across the OHDSI network the c-statistics ranged between 0.40-0.68.

**Interpretation:** The results show that the discriminative performance of the C-19 model is low for influenza cohorts, and even worse amongst COVID-19 patients in the US, Spain and South Korea. These results suggest that C-19 should not be used to aid decision making during the COVID-19 pandemic. Our findings highlight the importance of performing external validation across a range of settings, especially when a prediction model is being extrapolated to a different population. In the field of prediction, extensive validation is required to create appropriate trust in a model.

## Introduction

### Background and objectives

The novel severe acute respiratory syndrome coronavirus 2 (SARS-CoV-2) virus, also known as COVID-19, is quickly spreading throughout the world and burdening healthcare systems worldwide [1]. Numerous prediction models have started to be developed and released to the public to aid decision making during the pandemic [2]. Many of these models aim to inform people of their risk of developing severe outcomes due to COVID-19 [3-5]. A recent systematic review found all then-published models suffer from high risks of bias along with one or more limitations, including small datasets used to develop the models and lack of external validation.[2]

The COVID-19 vulnerability (C-19) index [5] is an example of a model developed to identify people susceptible to severe outcomes during COVID-19 infection. The model is potentially valuable because it aims to predict the hospitalization risk in the general population [2]. The model publication is currently available as a preprint and the model is publicly available at the website http://c19survey.closedloop.ai. The C-19 index aims to predict which patients will require hospitalization due to pneumonia (or proxies for pneumonia) within 3 months. The model was developed using retrospectively collected Medicare data (patients aged 65 or older) that do not contain COVID-19 patients. There is, however, no guarantee that a model trained on non-COVID-19 Medicare patients will perform similarly or even adequately in COVID-19 patients. Moreover, no external validation of the model was presented in the model development paper. Research has shown that there is high risk of bias for a model that lacks external validation [6]. In addition, it is recommended that knowledge of model reproducibility and transportability is assessed before a model is used clinically [7]. Models must be reliable as poor predictions can hurt decision making [2].

The Observational Health Data Science and Informatics (OHDSI) collaboration is a group of researchers collaborating to develop best practices for analyzing observational healthcare data [8]. OHDSI has developed a framework that enables timely validation of prediction models across a large number of datasets from around the globe [9]. The OHDSI network currently contains large COVID-19 cohorts from the US, Europe and Asia. In this study we aim to demonstrate the importance of performing external validation before we can trust a model’s predictions. As a case study we chose to investigate the predictive performance of the C-19 index when applied to COVID-19 data from across the world. This study can inform us about the suitability of utilizing the C-19 model to aid decision making during the COVID-19 pandemic.

## Methods

Three models were developed in the C-19 index publication [5]. The simplest model was a logistic regression with a limited number of predictors: age, sex, hospital usage, 11 comorbidities and their age interactions. The two other models were less parsimonious gradient boosting machines with more than 500 variables. Only one of these gradient boosting machine models was reported. Withholding a model makes it non-compliant with the Transparent Reporting of a multivariable prediction model for Individual Prognosis Or Diagnosis (TRIPOD) statement [10] and makes external validation impossible. In this paper we chose to evaluate the simple logistic regression model, recognizing that COVID-19 prediction models are urgently needed worldwide, and parsimonious models are more readily implemented across healthcare settings.

### Source of data

Electronic medical records (EMR) and administrative claims databases from primary care and secondary care containing patients from Australia, Japan, Netherlands, Spain, South Korea, and the US were analyzed in a distributed network, and are detailed in the **Supplementary Appendix, Table S1**. Five datasets contained COVID-19 cases and nine datasets did not. All datasets used in this paper were mapped into the OHDSI Observational Medical Outcomes Partnership Common Data Model (OMOP-CDM) [11]. The OMOP-CDM was developed to enable researchers with diverse datasets to have a standard database structure. This enables analysis code and software to be shared among researchers which facilitates external validation of prediction models. De-identified or pseudonymised data were obtained from routinely collected records from clinical practice. Analyses were performed using the following databases: AU-ePBRN (linked primary and secondary care from Australia); JMDC (Japanese claims); IPCI (primary care EMR from The Netherlands); SIDIAP (primary care EMR from Spain); AUSOM and HIRA (EMR and claims from South Korea); CCAE, ClinFormatics, MDCR, MDCD (US claims), Optum EHR, Veteran Affairs (VA), CUIMC and TRDW (EMRs for the US). All analyses were conducted locally in a distributed network, where analysis code was sent to participating sites and only aggregate summary statistics were returned, with no sharing of patient-level data between organizations.

### Consent to publish

Each site obtained institutional review board (IRB) approval for the study or used de-identified data, and therefore the study was determined not to be human subjects research. Informed consent was not necessary at any site.

### Participants

The purpose of the C-19 index is to identify which COVID-19 patients are more likely to require hospitalization due to severe complications. Therefore, we investigated the model performance when applied to patients at an outpatient (OP) or emergency room (ER) visit who have either i) COVID-19 positive test or diagnosis (in databases with COVID-19 data) or ii) influenza or influenza-like symptoms (in databases without COVID-19 data, as a proxy for COVID-19) and no recent prior symptoms or pneumonia. We chose this approach as it mimics the situation where patients first seek treatment or medical advice due to developing symptoms or testing positive for COVID-19 (or influenza).

For external validation in COVID-19 data we defined a cohort of patients presenting at an initial healthcare provider interaction during an OP/ER visit with COVID-19 disease. COVID-19 disease was identified by a diagnosis code for SARS-COV-2 or a positive test for the SARS-COV-2 virus that was recorded after January 1^st^ 2020. We required patients to be aged 18 or over, to have at least 365 days of observation time prior to the index date and to have no diagnosis of influenza, influenza-like symptoms, or pneumonia in the preceding 60 days.

For the influenza validation, we identified patients aged 18 or older with an OP/ER visit with influenza or influenza-like symptoms (i.e. fever and either cough, shortness of breath, myalgia, malaise, or fatigue), at least 365 days of prior observation, and no diagnosis of influenza, influenza-like symptoms, or pneumonia in the preceding 60 days.

### Outcome

The outcome was hospitalization with pneumonia on the index date (valid OP/ER visit) and within 30 days after index.

**Appendix A** contains the definitions for pneumonia, influenza, influenza-like symptoms and COVID-19 used in this study. The full details of the target population cohorts and outcomes used for validation can be found in the study package.

### Predictors

The predictors of the logistic regression version of the C-19 index are age in years, male sex, number of inpatient visits during the prior 12 months and indicator variables for various Clinical Classifications Software Refined (CCSR) categories. A table with the C-19 predictors and coefficients is presented in **Appendix B**. The CCSR categories used were pneumonia except that caused by tuberculosis, other and ill-defined heart disease, heart failure, acute rheumatic heart disease, coronary atherosclerosis and other heart disease, pulmonary heart disease, chronic rheumatic heart disease, diabetes mellitus with complication, diabetes mellitus without complication, chronic obstructive pulmonary disease and bronchiectasis, other specified and unspecified lower respiratory disease. Age interactions with each CCSR variable were also included as predictors. Each CCSR category corresponds to an aggregation of ICD-10 codes that belong to the category.

In the development data, if a patient had an ICD-10 code that was part of the CCSR ‘pneumonia except that caused by tuberculosis’ grouping during a specified time period prior to index their value for the predictor ‘pneumonia except that caused by tuberculosis’ was 1 and 0 otherwise. This was repeated for each CCSR predictor. Data in the OMOP-CDM do not use ICD-10 codes, but instead use Systematized Nomenclature of Medicine (SNOMED) codes. Therefore, to replicate the predictors in the OMOP-CDM data we needed to find the sets of SNOMED codes that correspond to each CCSR predictor. We accomplished this by finding the SNOMED equivalent of each ICD-10 code in a CCSR category.

The SNOMED groupings per CCSR category used by the OHDSI implementation of the C-19 are presented in **Appendix B**.

### Sample Size

We identified 41,381 patients with an OP/ER visit for COVID-19 in 2020: 1,985 patients from South Korea, 37,950 patients from Spain and 1,446 patients from the US. We also identified a total of 9,429,285 patients with an OP/ER visit for influenza or influenza-like symptoms in databases from 6 countries. The number of visits for influenza or influenza-like symptoms per database ranged between 2,793 to 3,146,801.

### Missing Data

The prediction models used a cohort design that includes any patient that satisfies the inclusion criteria. We did not exclude patients who are lost to follow-up during the 30-day period after the valid OP/ER visit.

### Statistical analysis methods

The model performance was evaluated using the standard discriminative metrics: area under the receiver operating characteristic (AUROC) curve (equivalent to the c-statistic) and area under the precision recall curve (AUPRC). The latter is a useful addition to the AUROC when assessing rare outcomes [12]. The calibration was determined by creating deciles based on the predicted risk and plotting the mean predicted risk versus the observed risk in each decile. If a model is well calibrated, then the mean predicted risk will be approximately equal to the observed risk for each decile.

We follow the TRIPOD statement guidelines [10] for reporting the model validation throughout this paper. For transparency an open source package for implementing the model on any OMOP-CDM data is available at https://github.com/ohdsi-studies/Covid19PredictionStudies/tree/master/CovidVulnerabilityIndex.

### Development vs. validation

The differences between the C-19 model development settings and the validation settings include a different target population and different datasets. Our validation design settings were chosen to mimic the COVID-19 situation when a clinician needs to decide whether to admit a patient. Importantly, we validated the C-19 model on COVID-19 patients.

The C-19 index was developed using a cohort design that entered adult patients into the cohort on 9/30/2016 and predicted whether they would be hospitalized for pneumonia or proxies (influenza, acute bronchitis, or other specified upper respiratory infections) in the following 3 months. Patients must have been in the data for 6 or more months and patients who left the database within 3 months of index and did not have death recorded were excluded. In our external validation we used a cohort design but entered adult patients into the cohort when they had an initial OP/ER visit for influenza (or COVID-19) rather than a fixed date and predicted hospitalization due to pneumonia in 30 days rather than 3 months. We excluded patients with influenza or pneumonia within the 60 days prior to index to restrict to initial visits. This mimics the situation during the COVID-19 pandemic where clinicians need to decide whether to hospitalize a patient initially presenting with COVID-19. We required 12 months of prior observation and did not exclude patients who left the database within 3 months of index.

The C-19 index was developed using a subset of patients from the Medicare database prior to the pandemic. This is a US claims database containing patients aged 65 or older. In this study we were able to externally evaluate the C-19 model on COVID-19 data, including adult patients under 65 years of age, from South Korea, Spain and the US.

## Results

### Online results

The complete results are available as an interactive app at:

http://evidence.ohdsi.org/C19validation

The characteristics of the MDCR data (same data source as the development data but different patient subset) and the HIRA, SIDIAP and VA data (COVID-19 patients) are displayed in **Table 1**. The characteristics for all datasets used in the study are available in **Appendix C**.

**Table 1:** Characteristics of patients at baseline in MDCR (database similar to development data) and the datasets with COVID-19 data. * indicates censoring due to a low cell count.

|  | MDCR |  | HIRA |  | SIDIAP |  | VA |  |
| --- | --- | --- | --- | --- | --- | --- | --- | --- |
| Predictor | Required Hosp | No Hosp | Required Hosp | No Hosp | Required Hosp | No Hosp | Required Hosp | No Hosp |
| Mean age in years | 80.92 | 76.41 | 65.53 | 45.09 | 63.28 | 49.61 | 69.64 | 58.41 |
| Mean number of inpatient visits in prior 365 days | 0.58 | 0.35 | 1.38 | 0.68 | * | * | 0.32 | 0.2 |
| MALE (%) | 52 | 45 | 56 | 46 | 59 | 43 | 95 | 80 |
| Fraction of patients with a history of each following condition (not including index): |  |  |  |  |  |  |  |  |
| Acute rheumatic heart disease | 0.00 | 0.00 | 0.00 | 0.00 | * | * | * | * |
| Chronic obstructive pulmonary disease and bronchiectasis | 0.43 | 0.25 | 0.38 | 0.21 | 0.06 | 0.03 | 0.27 | 0.2 |
| Chronic rheumatic heart disease | 0.03 | 0.02 | 0.00 | 0.00 | * | * | * | * |
| Coronary atherosclerosis and other heart disease | 0.19 | 0.15 | 0.21 | 0.09 | 0.02 | 0.01 | 0.17 | 0.1 |
| Diabetes mellitus with complication | 0.24 | 0.18 | 0.31 | 0.13 | 0.03 | 0.01 | 0.38 | 0.2 |
| Diabetes mellitus without complication | 0.38 | 0.32 | 0.43 | 0.20 | 0.13 | 0.05 | 0.50 | 0.3 |
| Heart failure | 0.37 | 0.20 | 0.20 | 0.07 | 0.02 | 0.01 | 0.23 | 0.1 |
| Other and ill-defined heart disease | 0.25 | 0.15 | 0.02 | 0.01 | 0.01 | 0.01 | 0.11 | 0.0 |
| Other specified and unspecified lower respiratory disease | 0.73 | 0.59 | 0.92 | 0.88 | 0.43 | 0.38 | 0.58 | 0.4 |
| Pneumonia (except caused tuberculosis) | 0.39 | 0.20 | 0.31 | 0.15 | 0.06 | 0.06 | 0.20 | 0.1 |
| Pulmonary heart disease | 0.09 | 0.04 | 0.00 | 0.00 | * | * | * | * |

### Model performance

When C-19 was transported to COVID-19 patients it achieved AUROCs between 0.36-0.56, full details are available in **Table 2**. The AUROC and calibration plots are presented in **Figure 1**. The internal discriminative performance of the C-19 index was with an AUROC of 0.73. When we validated the model on MDCR patients, but with our target population consisting of symptomatic influenza patients, the performance was 0.65, a significant drop from the 0.73 development performance. The AUROC performance when externally validated to other databases containing influenza patients ranged between 0.40-0.68. Full results are presented in **Table 3**, and AUROC and calibration plots are presented in **Appendix D**. As a sensitivity analysis we also validated the C-19 index on a target population consisting of patients with COVID-19 or symptoms during 2020, the results were similar and are presented in **Appendix E**.

**Table 2.** External validation of the C-19 model on COVID-19 data. * The 95% CI is reported when the outcome count is less than 1000.

| Database | Target Cohort | T size | O size (%) | AUROC (95% CI) | AUPRC |
| --- | --- | --- | --- | --- | --- |
| HIRA | OP/ER visit with COVID-19 positive record in 2020 and | 1,985 | 89 (4.48) | 0.56 (0.488-0.636) | 0.07 |
| SIDIAP | no symptoms in prior 60 | 37,950 | 1,223 (3.22) | 0.363* | 0.03 |
| VA | days | 1,446 | 149 (10.30) | 0.529 (0.473-0.584) | 0.14 |

OP – Outpatient; ER – Emergency room; T – Target population; O – outcome; CI – confidence interval; AUROC – area under the receiver operating characteristic curve; AUPRC: area under the precision recall curve.

**Table 3.**
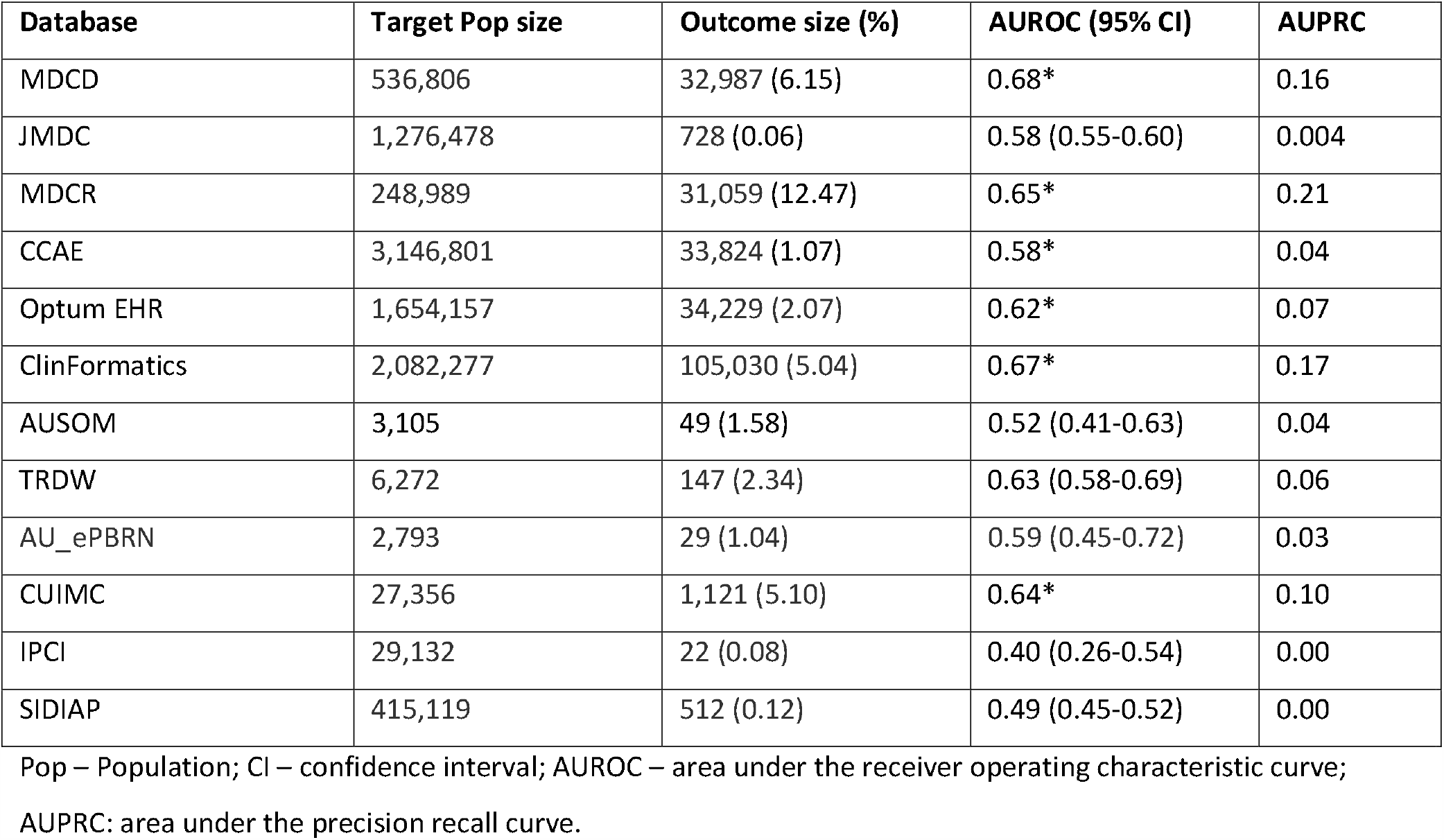
External validation of the C-19 model on influenza patient data. * The 95% CI is reported when the outcome count is less than 1000.

**Figure 1.**
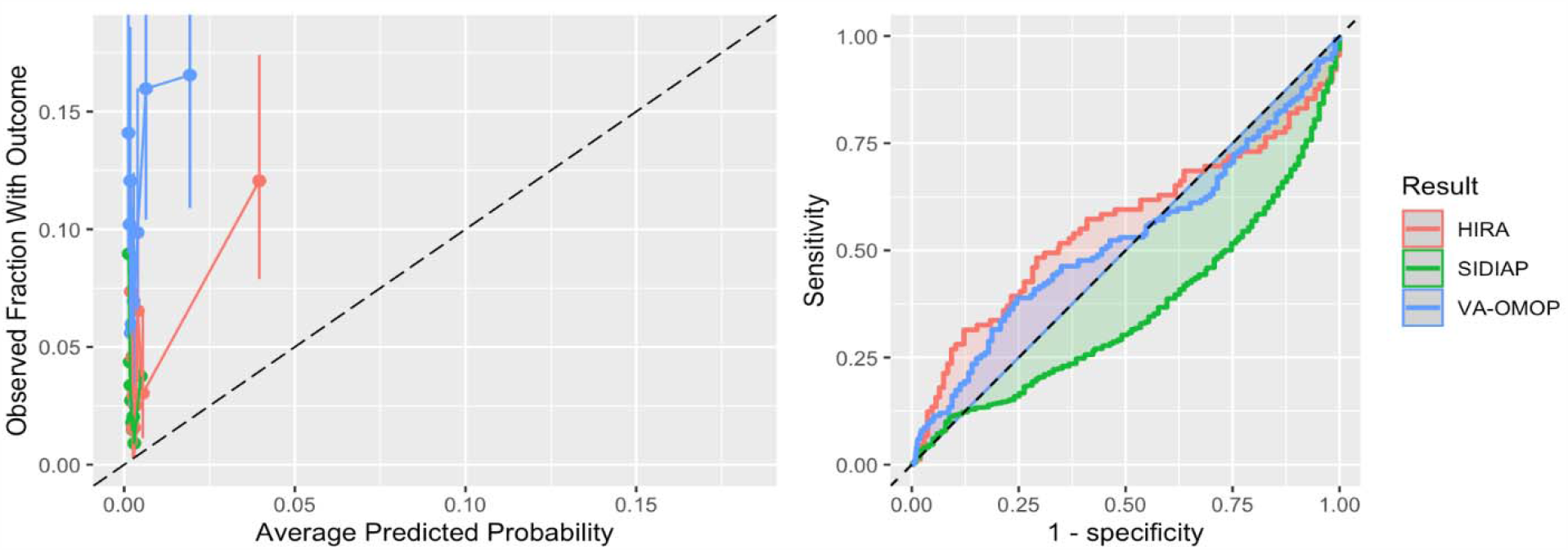
The ROC and calibration plots of C-19 for the three datasets with sufficient and suitable COVID-19 data.

## Discussion

The C-19 index is available online as a tool to predict severity in patients with COVID-19; while lacking validation for this population. Our validation across three datasets with sufficient COVID-19 data showed poor discriminative performance (AUROCs <0.6) and calibration. We observed similarly poor performance when validated across twelve datasets with influenza patients, with best AUROCs <0.70.

### Interpretation

The key finding of this study is the performance of the C-19 model when transported to COVID-19 patients. The model performance was poor (AUROCs 0.36-0.56) across the COVID-19 datasets. The performance was worse than random guessing in the SIDIAP data, which is consistent with the poor performance seen when applied to European patients with influenza. The calibration plots show that the C-19 index consistently underestimated risk in the COVID-19 patients.

The datasets used to perform the validation had very different patient populations. MDCR had the oldest patient population and many patients had comorbidities. Compared to MDCR, the CCAE and JMDC datasets presented healthier and younger patients (mean age around 40s) in the target population. While MDCD had younger patients these patients often had comorbidities (i.e. 20% these patients had COPD, 11% had heart failure and 17% has a history of pneumonia). The rate of hospitalization ranged greatly across the sites with values between 0.1% in JMDC and 12.4% in MDCR. The rate of the outcome in the dataset used to develop the C-19 index was 0.23%, much lower than in the MDCR data used to validate the model in this study. This is due to our study restricting to patients at the point they had an OP/ER visit due to influenza or COVID-19. Although five datasets contained COVID-19 patients, only four had sufficient data (VA, HIRA, SIDIAP and CUIMC) for external validation. The result of the C-19 when applied to COVID-19 patients in CUIMC was poor, <0.5 AUROC, however this dataset consisted mostly of hospitalized patients and therefore did not seem suitable for validating a model that predicts hospitalizations.

We chose a target population of symptomatic patients as this resembles the situation in which COVID-19 prediction models may be clinically implemented during the pandemic: clinicians likely would not admit asymptomatic patients. This suggests the internal C-19 AUROC estimate may be optimistic compared to if it were used in a realistic setting, due to the inclusion of many healthy patients. When applied to predict hospitalization in influenza patients across US data the performance ranged between 0.58-0.68. The performance was worse on the CCAE database with younger patients, likely due to age being a key predictor in the model. When the C-19 index was transported across non-US datasets the performance was poor to reasonable in the Australian and Asian data (0.52-0.64) and poor in the European data (0.4-0.49). The European data are extracted from general practice (GP) settings, but the C-19 model was developed using US claims data. Given the differences in clinical settings, it is not surprising that the performances were poor. This highlights that models often may not transport to different healthcare settings. The AUC of 0.36 when the C-19 model was validated in SIDIAP is worse than random guessing and inverting the predicted risk would lead to an AUC of 0.64. This may be a result of the C-19 including age interaction terms that resulted in the age coefficient being negative. Table 1 shows that in SIDIAP the model’s age interacting comorbidities are not as often recorded relative to the other databases. This may have resulted in younger patients being assigned higher risks than older patients in SIDIAP.

### Implication

The results provide extensive insight into the performance of the logistic regression C-19 index when used for COVID-19. The external validation uncovered that the logistic regression C-19 model is unreliable when predicting hospitalization risk for COVID-19 patients. Given this result, we do not recommend using the logistic regression C-19 index to aid decision making during the COVID-19 pandemic. The model did not appear to transport to COVID-19 patients, highlighting the importance of externally validating models, especially models whose target population differs from the development population.

There are numerous potential reasons why the logistic regression C-19 model failed to predict hospitalization due to pneumonia in the COVID-19 patients investigated. First reason may be due to the model being developed on patients aged 65 or older but applied to patients aged 18 or older. Age had a negative coefficient in the model, so this may have caused issues when the model was applied to younger patients. A second reason may be due to incorrect phenotyping for the predictors. We matched the SNOMED codes to the CCSR ICD-10 codes provided, but the predictors may require database specific phenotypes due to coding differences across datasets and healthcare settings. This may explain the poor performance in the European datasets that may record things differently than the US. A third reason is the study design:C-19 was developed to predict hospitalization from a set date in 2016 but we validated in a target cohort of symptomatic patients with an OP/ER visit as this more closely matches the clinical use case of the model. This means we are likely to have a sicker population where discrimination may be more difficult. A fourth potential reason is that the C-19 model was developed using data prior to 2017 but was validated on data from 2020: temporal changes and concept drift may negatively impact performance. Although we do not know the reason for the unreliability of the C-19 model on COVID-19 patients, we were able to quantify it by large-scale external validation across a network of datasets. In future work it would be beneficial to develop techniques that can identify reasons for poor external validation performance, as this may inform new best practices for model development.

This study highlights the importance of performing extensive external validation across different settings. During times of uncertainty, such as during pandemics, medical staff who are under pressure to make important decisions could benefit from implementing vetted prediction models. However, it is important to gain an unbiased and reliable evaluation of a model’s performance across numerous patient populations before the model is used. Internal validation can often be biased (e.g., the population used the develop the model does not match the intended target population) and provide optimistic performance estimates (e.g., a poor design or small dataset may result in overestimated discriminative performance). The approach used by the OHDSI collaboration enables efficient external validation of models across multiple datasets and this is a valuable resource when urgency is required.

### Limitations

A common issue when using observational healthcare data, especially across a network of databases, is the difficulty in developing phenotypes that are valid on all datasets. In this study we used predictor definitions given by the researchers who developed the model. However, these definitions may not transport across all the datasets and may account for some of the decrease in performance. We were also limited to validate the less complex C-19 model due to the large number of variables and lack of transparency for the more complex models.

## Conclusion

We have demonstrated the importance of implementing external validation in multiple datasets to determine the reliability of prediction models. We picked a newly developed model, the C-19 index, that aimed to predict which COVID-19 patients are at risk of severe complications due to the virus. The model reported an internal AUC of 0.73 but was deemed as having a high risk of potential bias [2]. The C-19 index addresses an important issue that could have greatly aided decision making during the COVID-19 pandemic, but its performance in COVID-19 patients was unknown. Our results show that the C-19 index performs poorly when applied to newly diagnosed COVID-19 patients in Asia, Europe and the US. Overall, we suggest that the model currently only be used to predict hospitalization due to pneumonia in older patients in the US. The results of this study demonstrate that internal validation performance should be considered optimistic estimates and a prediction model requires validation across multiple datasets in the target population where it will be used (or a close proxy), before it should be trusted.

## Data Availability

Data is not publicly available due to patient privacy concerns.

## Acknowledgements

We would like to acknowledge the patients who suffered from or died of this devastating disease, and their families and caregivers. We would also like to thank the healthcare professionals involved in the management of COVID-19 during these challenging times, from primary care to intensive care units.

The authors appreciate healthcare professionals dedicated to treating COVID-19 patients in Korea, and the Ministry of Health and Welfare and the Health Insurance Review & Assessment Service of Korea for sharing invaluable national health insurance claims data in a prompt manner.

## Author contributions

All authors made substantial contributions to the conception or design of the work; DPA and PBR led the acquisition of the data; JMR led the analysis; all authors were involved in the interpretation of data for the work; All authors have contributed to the drafting and revising critically the manuscript for important intellectual content; all authors have given final approval and agree to be accountable for all aspects of the work.

## Appendix

**Table S1.** Data sources formatted to the OMOP-CDM used in this research

| Database | Database Acronym | Country | Data type | Contains COVID-19 data? | Time period |
| --- | --- | --- | --- | --- | --- |
| IBM MarketScan® Medicare Supplemental Database | MDCR | US | Claims | No | 2000-2018 |
| Columbia University Irving Medical Center Data Warehouse | CUIMC | US | EMR | Yes | Influenza: 1990-2020<br>COVID-19: March-April 2020 |
| Health Insurance and Review Assessment | HIRA | South Korea | Claims | Yes | COVID-19: January to April 2020 |
| The Information System for Research in Primary Care | SIDIAP | Spain | GP and hospital admission EHRs linked | Yes | Influenza: 2006-2017<br>COVID-19: March 2020 |
| Tufts Research Data Warehouse | TRDW | US | EMR | Yes | Influenza: 2006-2020<br>COVID-19: March 2020 |
| Department of Veterans Affairs | VA | US | EHR | Yes | COVID: 1<sup>st</sup> March – 20 April 2020 |
| Ajou University School of Medicine Database | AUSOM | South Korea | EHR | No | 1996 - 2018 |
| Australian Electronic Practice based Research Network | AU-ePBRN | Australia | GP and hospital admission EHRs linked | No | 2012-2019 |
| IBM MarketScan® Commercial Database | CCAE | US | Claims | No | 2000-2018 |
| Integrated Primary Care Information | IPCI | Netherlands | GP | Yes | 2006-2020 |
| Japan Medical Data Center | JMDC | Japan | Claims | No | 2005-2018 |
| IBM MarketScan® Multi-State Medicaid Database | MDCD | US | Claims | No | 2006-2017 |
| Optum® De-Identified Clinformatics® Data Mart Database | ClinFormatics | US | Claims | No | 2000-2018 |
| Optum® de-identified Electronic Health Record Dataset | Optum EHR | US | Claims | No | 2006-2018 |

## Appendix A: Definitions used in the study

[excel file called exValConcepts]

## Appendix B: Predictor SNOMED codes

| Coefficient | Predictor | Snomed codes |
| --- | --- | --- |
| -5.49 | Intercept | NA |
| -0.014 | Age | NA |
| -0.212 | Male | 8507 |
| 0.186 | Number of inpatient visits in last year |  |
| -0.008 | Acute rheumatic heart disease | 313500,321578,321586,437897,4225090,4271684,4306131 |
| -0.269 | Chronic obstructive pulmonary disease and bronchiectasis | 255573,255841,256448,257004,257905,258780,259043,261325,261889,261895,440748,765431,4046986,4050731,4050732,4050733,4050734,4050961,4056405,4083395,4104506,4110048,4110049,4110056,4110635,4112826,4112828,4112836,4115044,4136683,4137525,4138392,4145496,4148124,4163244,4166508,4166517,4172303,4177944,4193588,4196712,4209097,4230358,4246105,4278831,4286497,4309350,4315386,37206130,40481763,42536541,42539089,42574216,42598711,43530693,44791725,44807895,45769389,46269701,46270376,46274062 |
| -0.014 | Chronic rheumatic heart disease | 313221,313500,315282,315295,317296,318776,319825,320429,320743,321578,321586,380126,435829,437897,439834,4006165,4040820,4057759,4062117,4078214,4108085,4108168,4117862,4122085,4143607,4154293,4155486,4156119,41695 |
|  |  | 68,4175807,4178585,4181949,4182110,4184497,4192358,4195677,4209569,4215619,4221806,4222174,4225090,4227150,4231452,4238914,4240289,4244437,4259295,4263730,4264740,4271684,4301373,4304541,4306131,4312621,4313828,4339544,43020465,43020466 |
| -0.529 | Coronary atherosclerosis and other heart disease | 315296,315830,315832,321318,761735,4068938,4078531,4108670,4116486,4119455,4119942,4127089,4155008,4155009,4155963,4161456,4161457,4161973,4161974,4184827,4198141,4201629,4231426,4262446,4264145,4310270,4324893,35615052,35615053,36712982,36712983,36712984,37209632,43531588 |
| -0.273 | Diabetes mellitus with complication | 192279,200687,201530,201531,318712,321822,376065,376112,376114,376683,376979,377552,377821,378743,380096,380097,435216,439770,442793,443592,443727,443729,443730,443731,443732,443733,443734,443735,443767,760977,760978,760979,760980,760989,761048,761050,761051,761053,761062,761063,765373,765375,765533,765650,4007943,4009303,4016047,4023792,4027121,4029420,4029423,4030664,4033942,4034964,4044391,4044392,4044393,4046332,4048028,4048029,4054812,4061725,4063569,4065354,4082346,4082347,4082348,4087682,4095288,4099216,4099652,4101478,4101887,4101892,4102176,4105016,4105172,4105173,4105639,4114426,4114427,4128221,4129225,4129520,4131117,4131908,4137220,4140466,4142579,4143689,4143857,4145542,4147406,4147504,4147577,4147719,4151453,4151946,4159742,4161670,4161671,4162095,4162239,4164174,4164175,4164176,4164632,4169240,4171406,4174977,4175440,4176925,4177050,4189418,4191611,4192852,4194970,4195043,4195044,4195045,4195498,4199039,4200875,4206115,4209538,4210128,4210129,4210872,4210874,4212441,4215719,4215961,4218499,4221344,4221487,4221495,4221933,4221962,4222415,4222553,4222687,4222876,4223303,4223463,4223734,4223739,4224254,4224419,4224709,4224879,4225055,4225656,4226121,4226238,4226354,4226798,4227210,4227657,4228112,4228443,4234742,4235260,4242528,4243625,4247107,4252356,4255399,4255400,4255401,4262282,4263090,4265913,4266041,4266042,4266637,4269870,4269871,4270049,4290822,4290823,4295011,4304701,4307319,4307799,4311708,4321756,4334884,4336000,4338900,4338901,35625717,35625718,35625719,35625722,35625723,35625724,35626036,35626037,35626038,35626039,35626041,35626042,35626043,35626044,35626046,35626047,35626067,35626068,35626069,35626070,35626071,35626072,35626087,35626088,35626761,35626762,35626763,35626764,35626765,35626904,35626905,36674199,36674200,36674651,36674652,36674752,36674753,36674765,36674766,36684827,36712670,36712686,36712687,36713094,36714116,36715571,36716853,36717156,37016179,37016180,37016348,37016349,37016350,37016353,37016354,37016355,37016356,37016357,37016358,37016767,37016768,37017221,37017429,37017430,37017431,37017432,37018566,37018728,37018912,37109305,37110068,37110593,37396268,40480000,40480031,40482458,42535539,42536400,42536603,42536604,42536605,42538169,42538715,43530656,43530660,43530685,43530689,43530690,43531559,43531562,43531563,43531564,43531565,43531566,43531577,43531578,43531588,43531597,43531608,43531616,43531651,43531653,44789318,44789319,44805212,44805628,44809809,45757065,45757073,45757074,45757075,45757255,45757266,45757277,45757278,45757280,45757362,45757363,45757392,45757393,45757432,45757435,45757444,45757445,45757446,45757447,45757449,45757450,45757499,45757507,45757535,45757604,45763582,45763583,45763584,45763585,45766963,45769828,45769829,45769830,45769832,45769833,45769834,45769835,45769836,45769837,45769872,45769873,45769875,45769876,45769888,45769889,45769890,45769891,45769892,45769894,45769901,45769902,45769903,45769904,45769905,45769906,45770830,45770831,45770832,45770880,45770881,45770883,45770902,45770928,45771064,45771067,45771068,45771072,45771075,45771533,45772019,45772060,45772914,45773064,45773567,45773576,45773688,46269764,46274058 |
| -0.496 | Diabetes mellitus without complication | 443412,4008576,4193704,45757474 |
| -0.271 | Heart failure | 312927,314378,315295,316139,316994,319835,439694,439696,439698,439846,442310,443580,443587,444031,444101,762002,762003,764871,764872,764873,764874,764876,764877,4004279,4009047,4014159,4023479,4030258,4071869,4079296,4079695,4103448,4108244,4108245,4111554,4124705,4134890,4138307,4139864,4141124,4142561,4172864,4177493,4184497,4185565,4193236,4195785,4195892,4199500,4205558,4206009,4215446,4215802,4229440,4233224,4233424,4242669,4259490,4264636,4267800,4273632,4284562,4307356,4311437,4327205,35615055,36712927,36712928,36712929,36713488,36716182,36716748,36717359,37110330,40479192,40479576,40480602,40480603,40481042,40481043,40482857, |
|  |  | 40486933,42598803,43020421,43020657,43021735,43021736,43021825,43021826,43021840,43021841,43021842,43022054,43022068,43530642,43530643,43530961,44782428,44782655,44782713,44782718,44782719,44782728,44782733,44784345,44784442,45766164,45766165,45766166,45766167,45766964,45773075 |
| -0.111 | Other and ill-defined heart disease | 4131824,43021898,40483752,4317287,4069185,42536628,40489421,43022035,4108352,4237062,4100397,4216844,35615119,40483223,40481472,36712751,40487039,4120089,4068741,43020564,43021610,43021955,42594384,42594383,42536629,4173820,4119953,35622329,438171,43021897,4273462,4100132,42599748,4170062,4175580,43021066,37109910,4101319,4182190,4236169,43020636,43020927,4119606,36712752,4033322,36716866,4321717,4100871,40479589,43021064,43020641,4119462,42536642,43021734,36712838,40487573,36712985,42534988,321320,42536633,43020582,42537536,4108220,4108219,4108722,43021065,438168,316427,4102852,4148905,43020889,43021891,4108950,314658,432937,4141491 |
| -0.02 | Other specified and unspecified lower respiratory disease | 4027553 plus all descendants |
| 0.117 | Pneumonia (except caused tuberculosis) | 252351,252548,252655,252949,253235,253790,254066,254561,254677,255084,255735,255848,256722,256723,257315,257908,258061,258180,258333,258354,258785,259048,259852,259992,260028,260041,260430,260754,261053,261324,261326,436145,437313,439857,440431,442637,443410,759815,759816,759817,759818,759821,763011,763012,4021760,4025165,4044215,4045227,4046011,4048052,4048147,4048148,4048149,4048517,4048518,4048519,4049965,4050872,4050874,4051333,4051334,4051335,4051337,4051338,4051339,4052546,4052547,4052548,4070540,4071610,4071611,4080435,4080753,4080883,4082065,4084973,4102253,4110039,4110506,4110507,4110509,4110510,4111119,4112655,4112820,4112822,4114030,4114031,4116487,4116488,4117114,4119431,4119436,4119795,4124539,4133224,4135197,4137435,4138244,4138769,4140134,4141619,4143092,4144107,4145369,4148529,4153356,4166072,4169796,4174308,4174309,4175598,4177385,4186568,4190647,4193964,4195014,4195452,4200891,4203846,4204819,4205578,4212120,4215807,4221503,4221767,4222062,4223032,4225318,4228277,4233319,4236311,4240452,4245006,4245499,4248029,4248154,4248807,4256236,4256894,4267135,4273378,4274802,4274981,4276663,4280213,4284985,4293463,4294404,4299862,4308451,4309106,4310964,4311555,4322625,4327820,4334649,4341520,4345215,4345699,35622404,36676238,36714118,37016927,37017277,37017278,37019058,37110291,37110292,37116366,37119233,37394479,40479642,40480033,40481335,40481839,40482061,42572644,42572881,42573020,42573178,42573179,42573181,42573218,42573349,42593423,42598655,42598908,42598979,42598991,42599060,42599199,42600053,42600167,43020558,44782989,45757206,45757250,45757644,45763749,45763750,45763751,45763752,45767051,45768914,45768960,45768961,45768997,45768998,45769390,45769809,45770900,45771022,46269693,46269707,46269708,46269709,46269710,46269711,46269712,46269713,46269714,46269715,46269716,46269717,46269718,46269719,46269720,46269721,46269722,46269723,46269724,46269725,46269726,46269954,46270027,46270121,46270318,46270510,46274035 |
| -0.005 | Pulmonary heart disease | 312927,315831,317000,433783,441593,4013643,4108610,4119611,4121462,4121620,4124831,4149211,4167085,4195892,4284110,35615055,36715093,40482858,40493243,42536630,42536631,44782560,44782561,44782562,44783618,44783619,44783620,44783621,44783622,44783623,44783624,44783625,44783626,45766142 |
| 0.003 | Acute rheumatic heart disease X Age | See above |
| 0.013 | Chronic obstructive | See above |
|  | pulmonary disease and bronchiectasis<br>X Age |  |
| -0.001 | Chronic rheumatic heart disease<br>X Age | See above |
| 0.011 | Coronary atherosclerosis and other heart disease<br>X Age | See above |
| 0.007 | Diabetes mellitus with complication<br>X Age | See above |
| 0.009 | Diabetes mellitus without complication<br>X Age | See above |
| 0.009 | Heart failure<br>X Age | See above |
| 0.003 | Other and ill-defined heart disease<br>X Age | See above |
| 0.006 | Other specified and unspecified lower respiratory disease<br>X Age | See above |
| 0.01 | Pneumonia (except caused tuberculosis)<br>X Age | See above |
| 0 | Pulmonary heart disease<br>X Age | See above |

## Appendix C: Characteristics for all databases

Excel sheet: cv19indexCovs.csv

## Appendix D: ROC and calibration plots

Full results are available from http://evidence.ohdsi.org/C19validation

Plots using Target population of patients with influenza or influenza-like symptoms

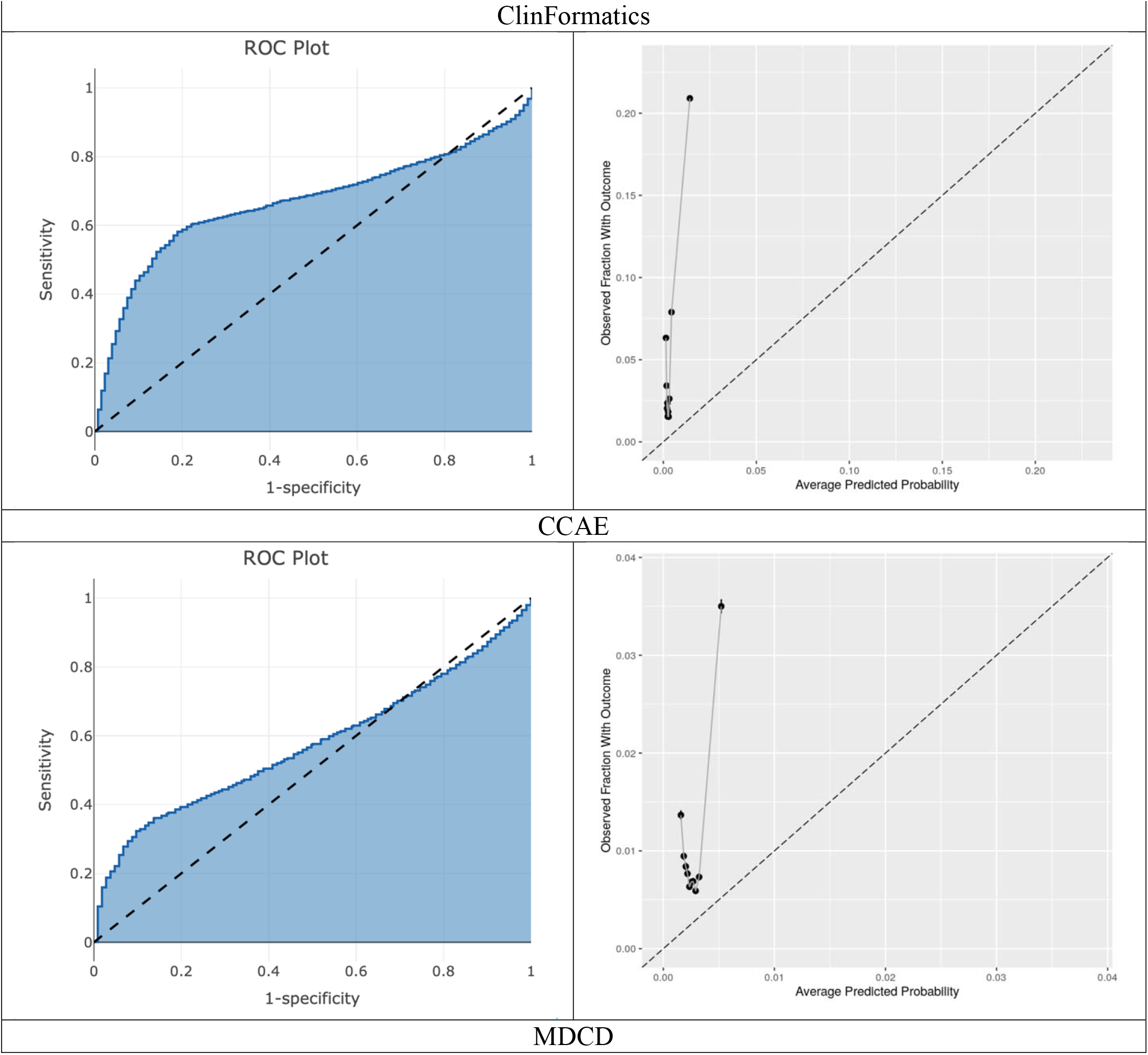
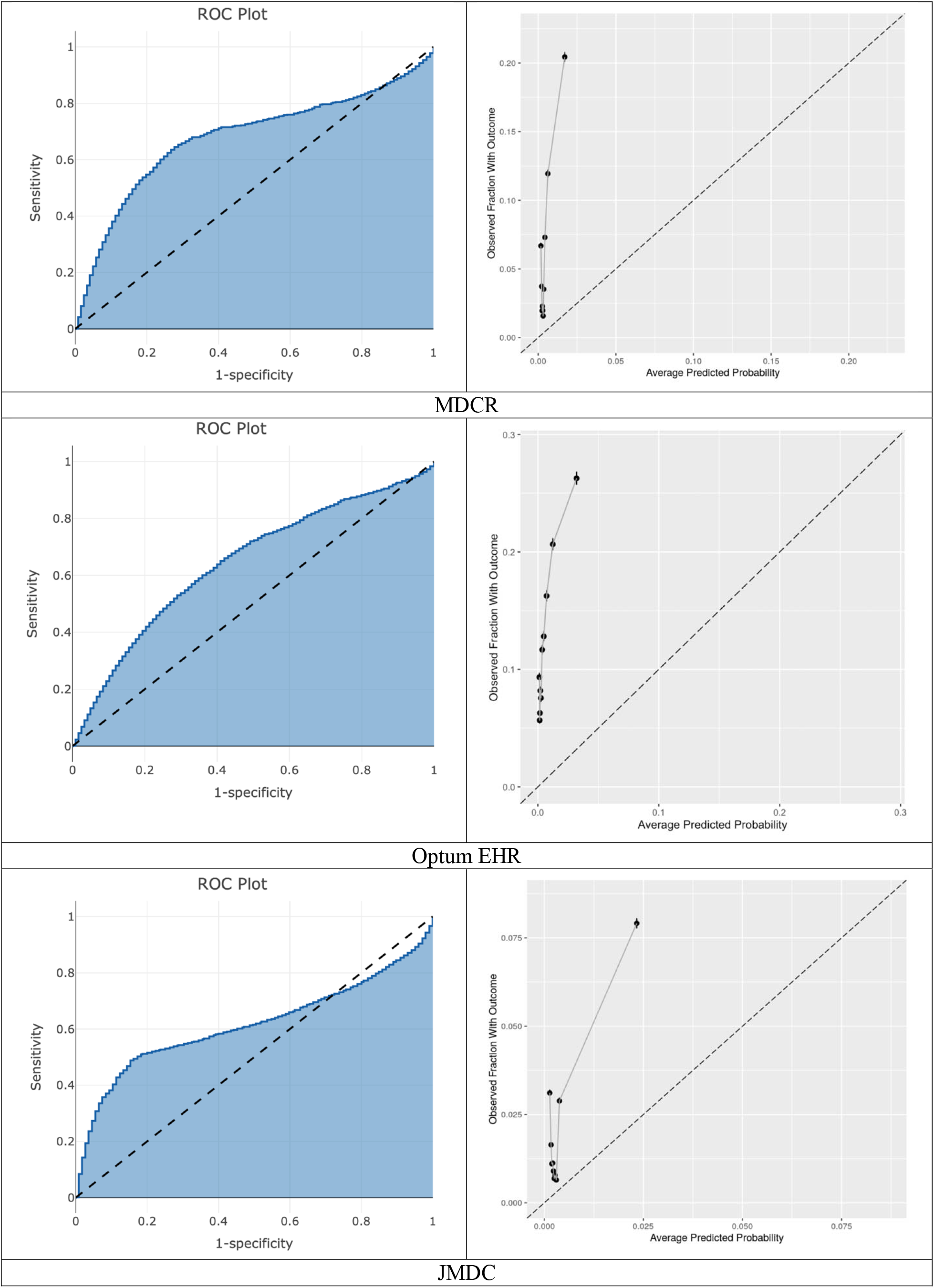
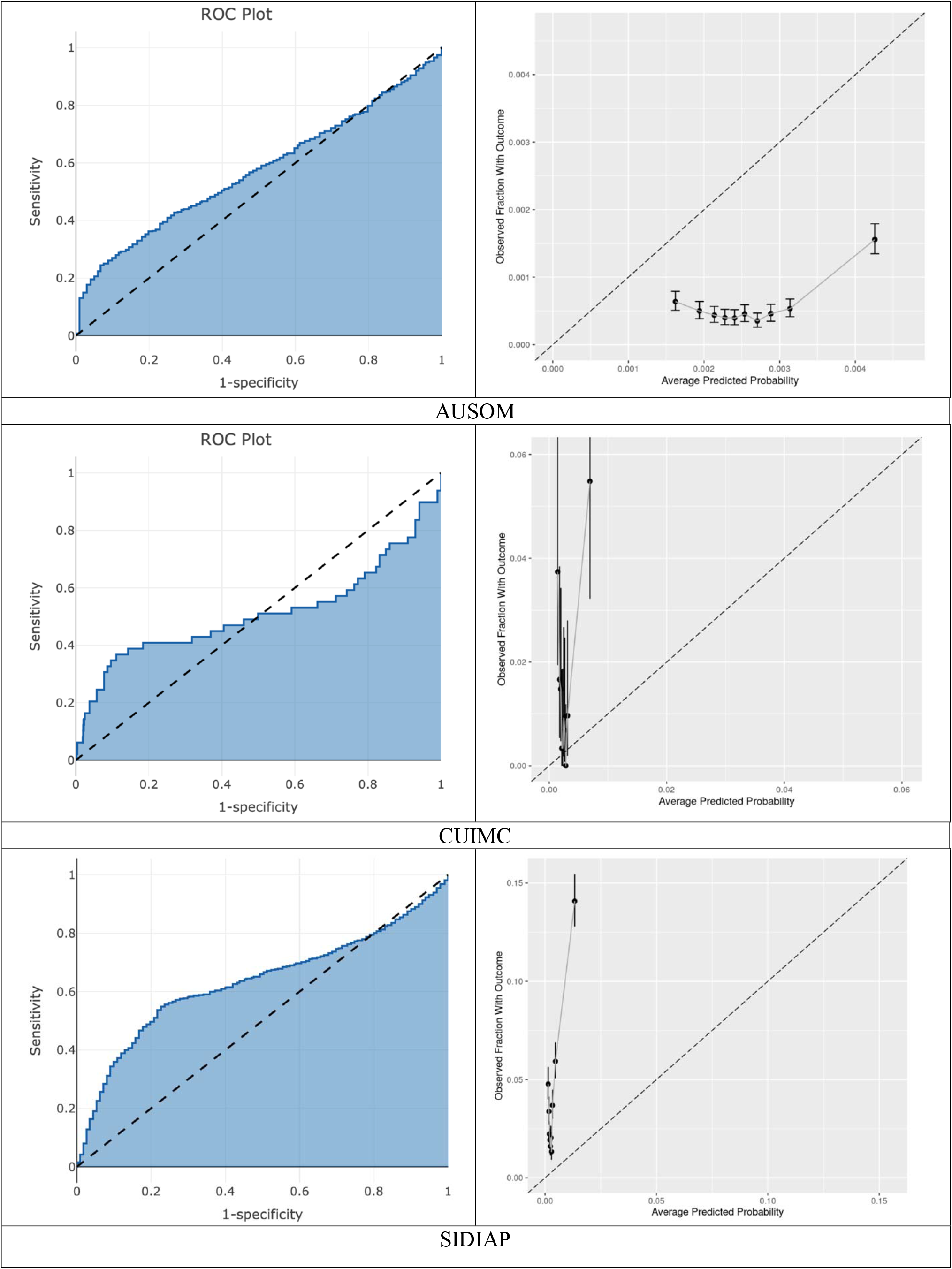
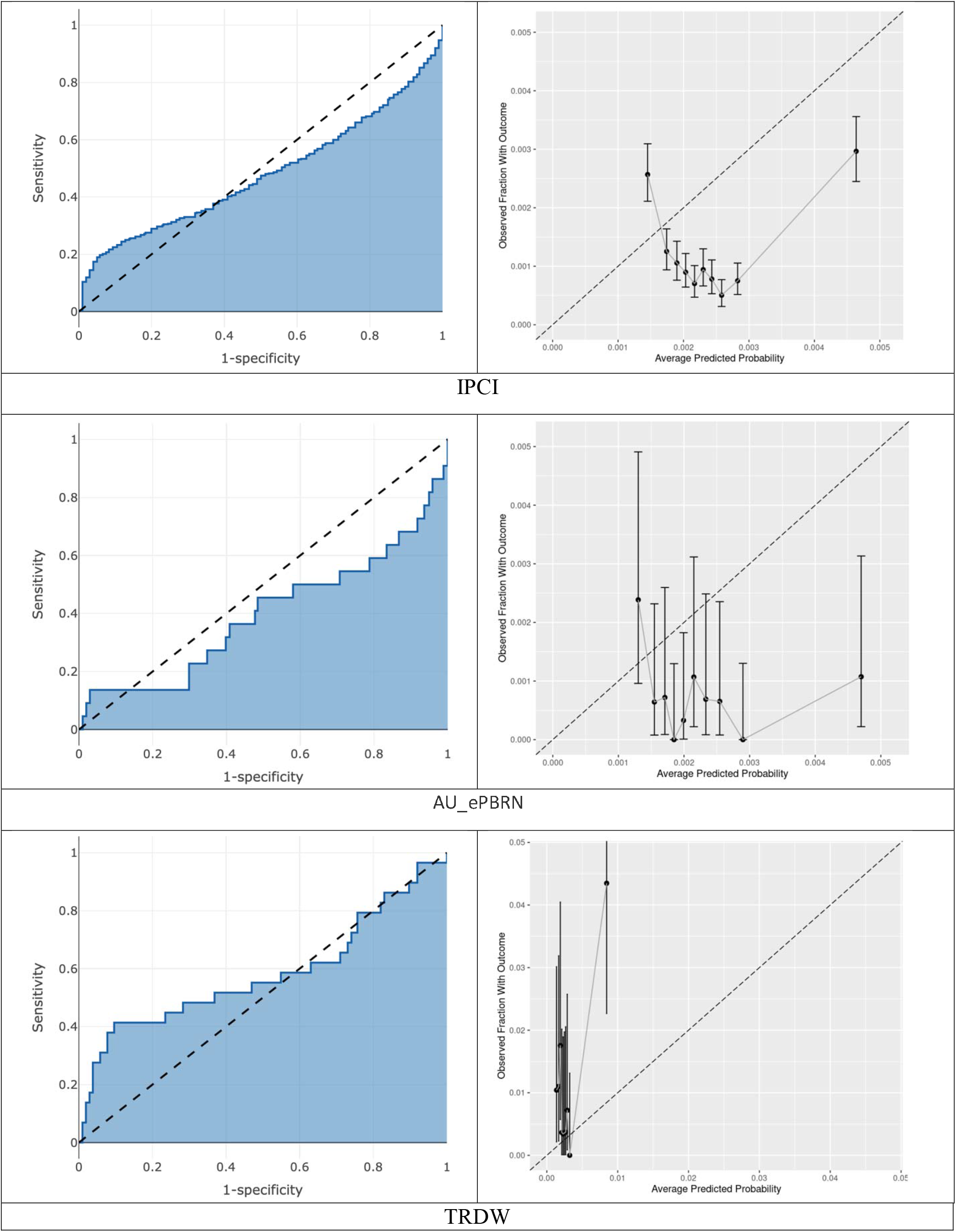
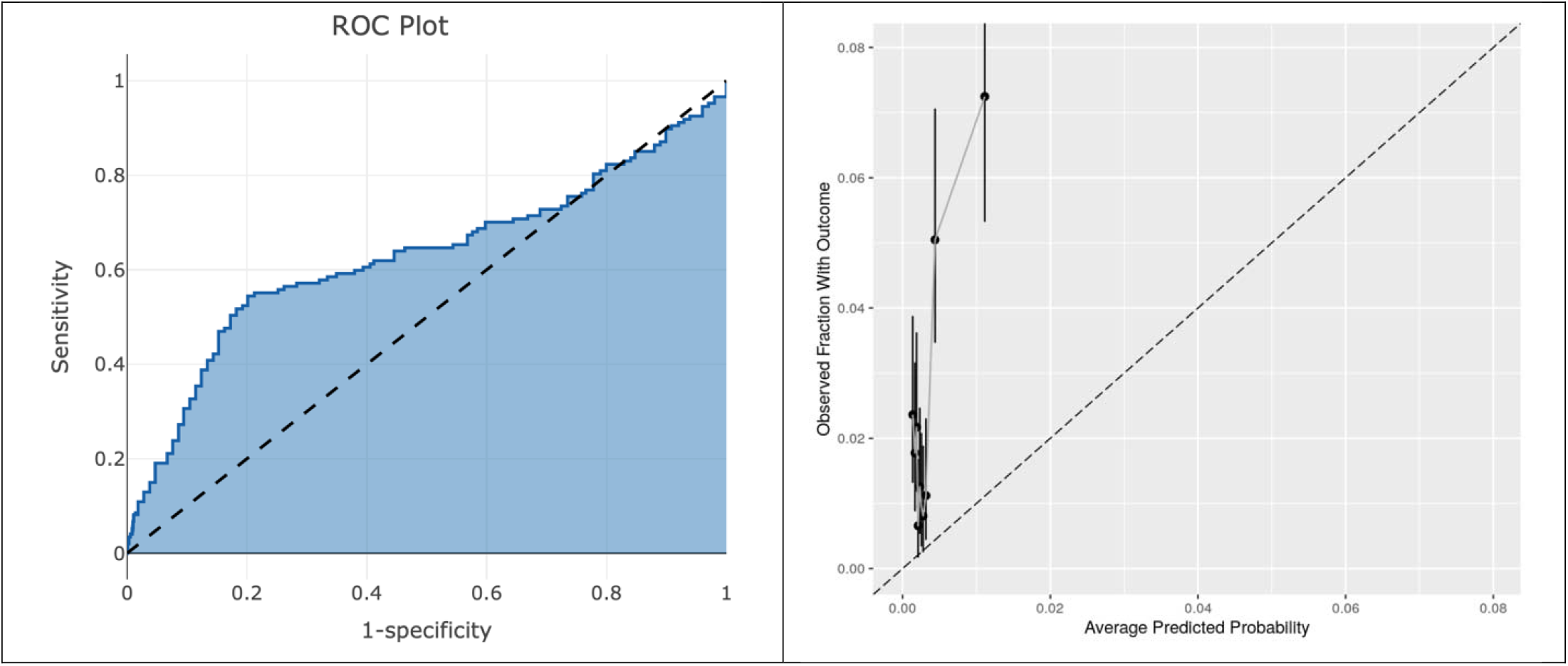

## Appendix E

| Database | Sensitivity Target Population | Target Population size | Outcome size (%) | AUROC | AUPRC |
| --- | --- | --- | --- | --- | --- |
| HIRA | COVID-19 positive test or symptoms in 2020 | 47,594 | 2,463 (5.18) | 0.64 | 0.1 |
| HIRA | COVID-19 positive test in 2020 | 1,985 | 89 (4.48) | 0.56 | 0.07 |
| TRDW | COVID-19 positive test or symptoms in 2020 | 285 | 5 (1.75) | 0.74 | 0.04 |
| SIDIAP | COVID-19 positive test or symptoms in 2020 | 38,254 | 1,229 (3.21) | 0.366 | 0.03 |
| SIDIAP | COVID-19 positive test in 2020 | 37,950 | 1,223 (3.22) | 0.363 | 0.03 |
| VA | COVID-19 positive test or symptoms in 2020 | 5,990 | 486 (8.11) | 0.627 | 0.15 |
| VA | COVID-19 positive test in 2020 | 1,446 | 149 (10.30) | 0.529 | 0.14 |

## References

1. Remuzzi A, Remuzzi G. COVID-19 and Italy: what next?. The Lancet. 2020 Mar 13.

2. Wynants, L., Van Calster, B., Bonten, M.M., Collins, G.S., Debray, T.P., De Vos, M., Haller, M.C., Heinze, G., Moons, K.G., Riley, R.D. and Schuit, E., 2020. Prediction models for diagnosis and prognosis of covid-19 infection: systematic review and critical appraisal. bmj, 369

3. Zhang H, Wang X, Fu Z, Luo M, Zhang Z, Zhang K, He Y, Wan D, Zhang L, Wang J, Yan X. Potential Factors for Prediction of Disease Severity of COVID-19 Patients.

4. Lu J, Hu S, Fan R, et al. ACP risk grade: a simple mortality index for patients with confirmed or suspected severe acute respiratory syndrome coronavirus 2 disease (COVID-19) during the early stage of outbreak in Wuhan, China. medRxiv [Preprint] 2020.doi:10.1101/2020.02.20.20025510

5. DeCaprio D, Gartner J, Burgess T, Kothari S, Sayed S. Building a COVID-19 Vulnerability Index. arXiv preprint arXiv:2003.07347. 2020 Mar 16.

6. Wolff RF, Moons KG, Riley RD, Whiting PF, Westwood M, Collins GS, Reitsma JB, Kleijnen J, Mallett S. PROBAST: a tool to assess the risk of bias and applicability of prediction model studies. Annals of internal medicine. 2019 Jan 1;170(1):51–8.

7. Van Calster - Van Calster, B., Wynants, L., Timmerman, D., Steyerberg, EW. and Collins, GS., Predictive analytics in health care: how can we know it works?. Journal of the American Medical Informatics Association 2019;26(12):1651–1654.

8. Hripcsak G, Duke JD, Shah NH et al. Observational health data sciences and informatics (OHDSI): opportunities for observational researchers. Stud Health Technol Inform 2015;216:574–578.

9. Reps, J.M., Williams, R.D., You, S.C., Falconer, T., Minty, E., Callahan, A., Ryan, P.B., Park, R.W., Lim, H.S. and Rijnbeek, P., 2020. Feasibility and evaluation of a large-scale external validation approach for patient-level prediction in an international data network: validation of models predicting stroke in female patients newly diagnosed with atrial fibrillation. BMC Medical Research Methodology, 20, pp.1–10.

10. Collins GS, Reitsma JB, Altman DG, Moons KG. Transparent reporting of a multivariable prediction model for individual prognosis or diagnosis (TRIPOD): the TRIPOD statement. British Journal of Surgery. 2015 Feb;102(3):148–58.

11. Erica A Voss, Rupa Makadia, Amy Matcho, Qianli Ma, Chris Knoll, Martijn Schuemie, Frank J DeFalco, Ajit Londhe, Vivienne Zhu, Patrick B Ryan, Feasibility and utility of applications of the common data model to multiple, disparate observational health databases, Journal of the American Medical Informatics Association, Volume 22, Issue 3, May 2015, Pages 553–564

12. Saito T, Rehmsmeier M. The precision-recall plot is more informative than the ROC plot when evaluating binary classifiers on imbalanced datasets. PLoS One. 2015;10(3):e0118432. Published 2015 Mar 4. doi:10.1371/journal.pone.0118432

